# A very simple model to account for the rapid rise of the British variant of SARS-CoV-2 in several countries and the world

**DOI:** 10.1101/2021.04.13.21254841

**Authors:** Hugo Fort

## Abstract

Since its first detection in the UK in September 2020, a highly contagious version of the coronavirus, called the British variant or B.1.1.7 SARS-CoV-2 virus lineage, is rapidly spreading across several countries and becoming the dominant strain in the outbreak.

Here it is shown that a very simple evolutionary model, when including the latest available data from March 2021, can fit the observed change in frequency of B.1.1.7 for several countries, regions of countries and the whole world with a single parameter which is almost universal.

---

In September 2020 a new variant of the SARS-CoV-2 virus, known as lineage B.1.1.7 (aka 20I/501Y.V1, Variant of Concern 202012/01, British variant), was detected in the UK and it quickly displaced the other variants of this virus in several countries (Hodcroft 2021).

Previous estimates of B.1.1.7 transmissibility have varied across different studies. For example: a preprint by Davis et al. (2020) reported that the variant was 56% (50%–74%) more transmissible than other variants across three regions in England (East of England, South East of England, and London), while another article concluded that it was 75% (70%–80%) more transmissible in the UK between October and November 2020 (Leung et al. 2021).

Here a very simple evolutionary model is proposed for describing the change in frequency of B.1.1.7. This model distinguishes the Brit variant B1.1.7 from all the others (treated just as one ‘mean’ lineage). If we call *x* the fraction of the B1.1.7 variant a simple equation to model evolution by natural selection is given by:

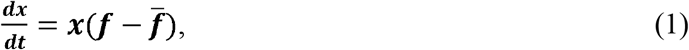

where ***f*** is the fitness of this lineage ***f*** relative to the others (which are set to 1) and 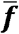 is the mean fitness over all the variants, i.e.

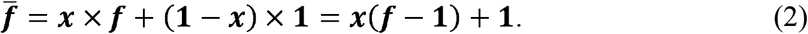

Substituting (2) into (1) we obtain:

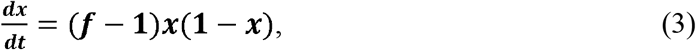

i.e. a logistic equation with a growth rate of *f*−1. The solution of (3) is (Fort 2021; 2020):

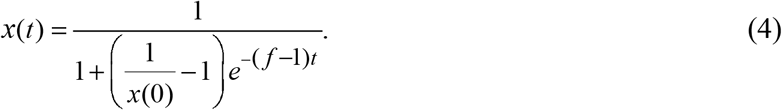

For long times, if *f* > 1, *x* will converge to an asymptotic value *x* =1.

Then, Eq.(4) is compared against empirical data for those countries such that a minimum statistics of 500 sequences in each measurement were included (Table 1). That is, countries are considered if they have at least *N*_*min*_ = 500 sequences summing, at each time, all the variants that were tracked since the first time the B.1.1.7 lineage was reported in each country (Hodcroft 2021).

**Table 1.** Data for the countries included in this study. *N*_*min*_ stands for the minimum total number of sequences (not cases) measured (see text).

| *Country* | *$N_{min}$ across weeks* | *Period* | *Relative fitness $f$* |
| --- | --- | --- | --- |
| Denmark<sup>1</sup> | 519 | 2020-02-26 to 2021-03-21 | 1.51 |
| Netherlands<sup>2</sup> | 500 | 2020-07-12 to 2021-3-8 | 1.33 |
| Switzerland<sup>2</sup> | 883 | 2020-07-12 to 2021-3-8 | 1.50 |
| UK<sup>2</sup> | 8217 | 2020-16-10 to 2021-3-8 | 1.48 |

It turns out that this formula, when including the latest available data from March 2021, can fit the observed change in frequency of B.1.1.7 for several countries, regions of countries and the whole world with a single parameter which is almost universal.

Fig. 1 shows how this fraction *x* has grown in the four countries of Table 1 and the comparison with the values predicted by Eq. (4). Notice that this formula in general fits very well with the data. Furthermore, in particular the data corresponding to Denmark and UK yield a clean sigmoid curve when plotted vs. time. This would be expected for a growth experiment (under controlled laboratory conditions), but not so much for this kind of measurements involving factors not controlled and different kind of heterogeneities.

**Figure 1:**
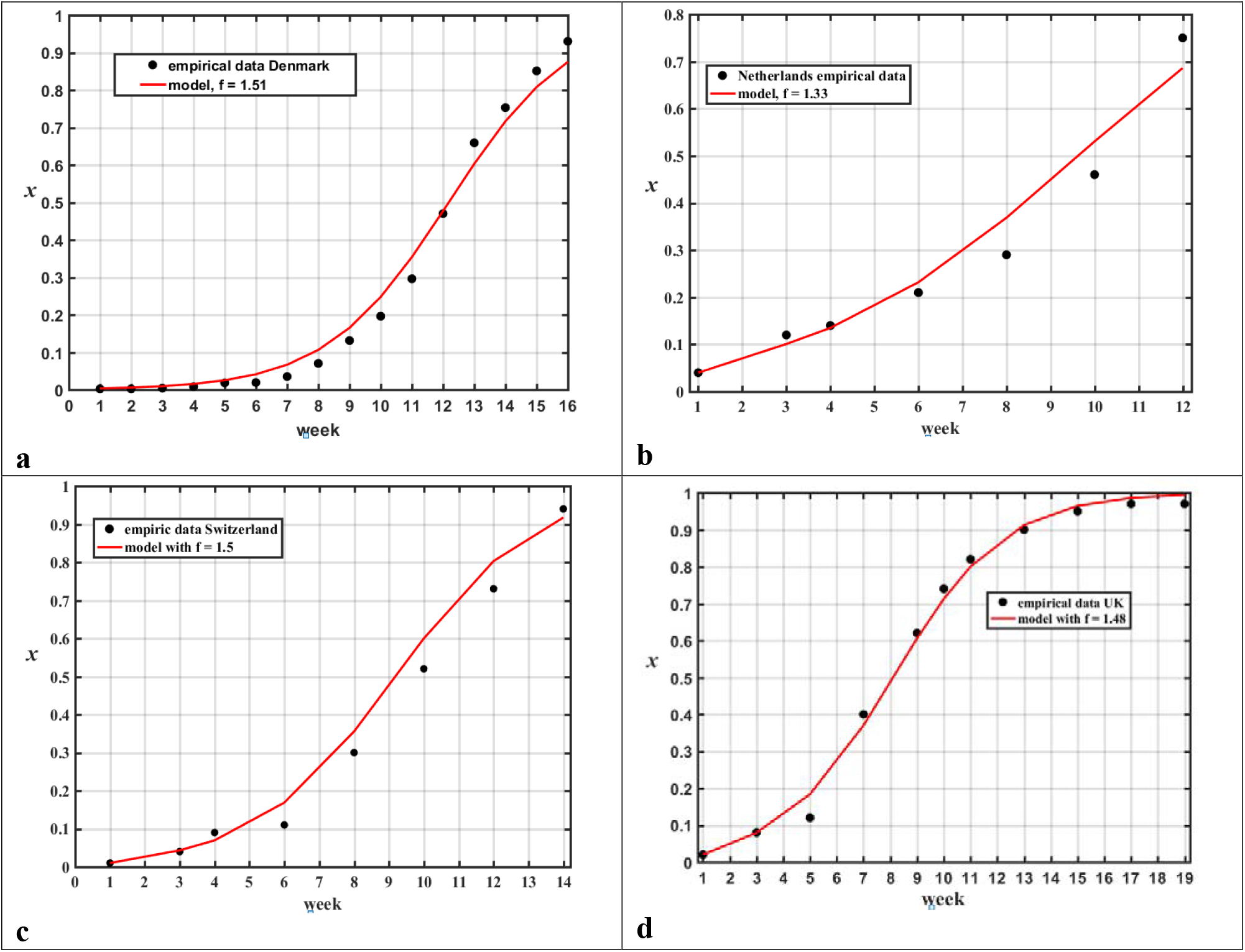
Change in the frequency of B.1.1.7 in the four countries of Table 1; empirical data (black circles) and formula (4) (solid line). (a) Denmark from November 2020 to 14 March 2021. (b) Netherlands from 7 December 2020 to 8 March 2021. (c) Switzerland from 7 December 2020 to 8 March 2021. (d) UK from 26 October 2020 to 8 March 2021

In the case of UK the frequency of B1.1.7 almost reached its asymptotic value *x* =1.

Interestingly, the relative fitness parameter for this British variant for the four countries, except Netherlands, is very similar ≈1.5. Additionally, the values of *f* =1.51 and *f* =1.50 for, respectively, Denmark and Switzerland are in agreement with a reported 52% higher transmissibility when compared to the wildtype in Denmark and a 51% higher transmissibility when compared to the wildtype in Switzerland (ISPM 2021). This suggests that it is right to interpret *f* as the relative fitness to the local wildtype.

It seems interesting to find out if the model still works for smaller spatial scales, for instance across different regions of a country. In fact, in many countries, sampling may not be equal across the country: samples may only cover one area or certain areas (Hodcroft 2021). Fig.2 shows the data for the five different regions of Denmark. Notice that, except for Nordjylland, the model fits the data well.

**Figure 2:**
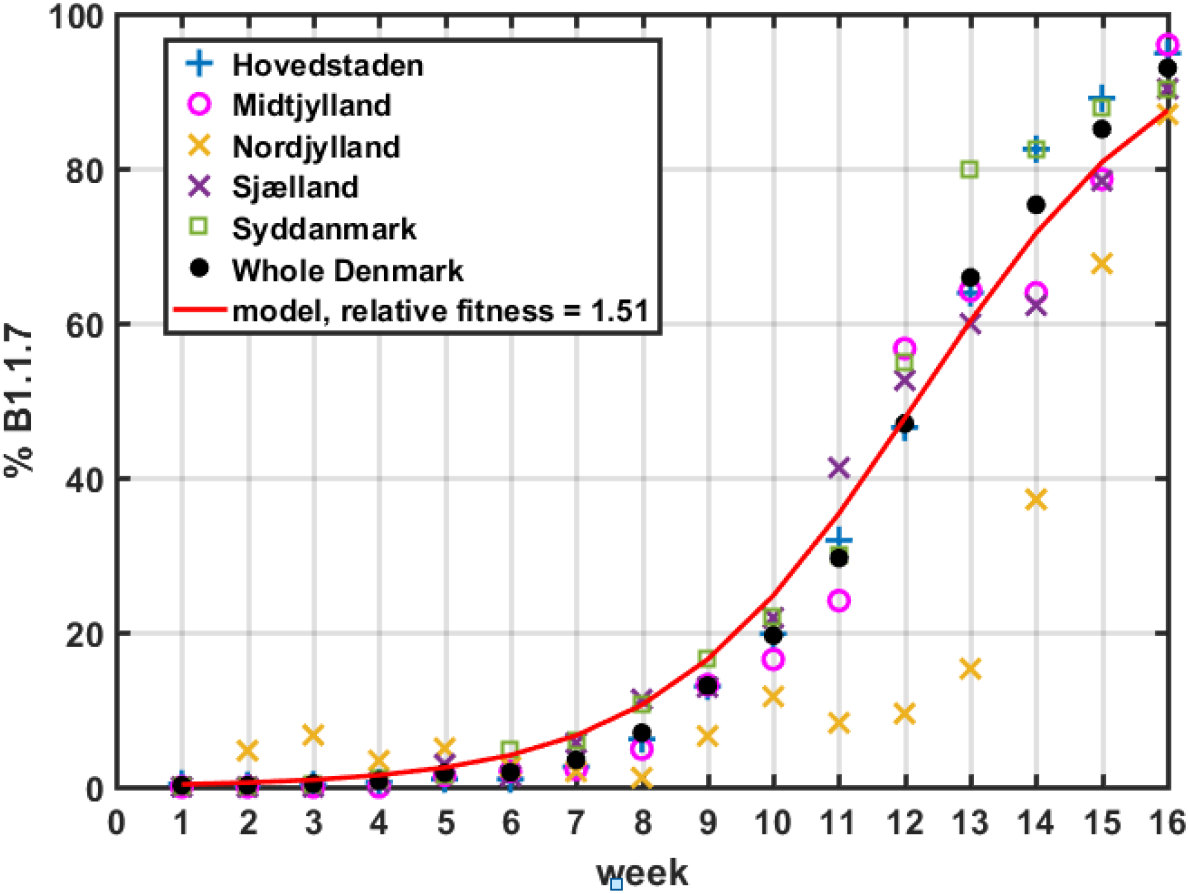
Change of the frequency of B.1.1.7 for different regions of Denmark as a percentage, empirical data (symbols) and formula (4) (red line).

Finally, let us see now the application of the model to the maximum spatial scale, i.e. the whole world. Fig. 3 shows the empirical data for the world (B.1.1.7 Lineage Report 2021) and the fit provided by Eq.(4) yields again a remarkable fit with the data for *f* =1.54.

**Figure 3:**
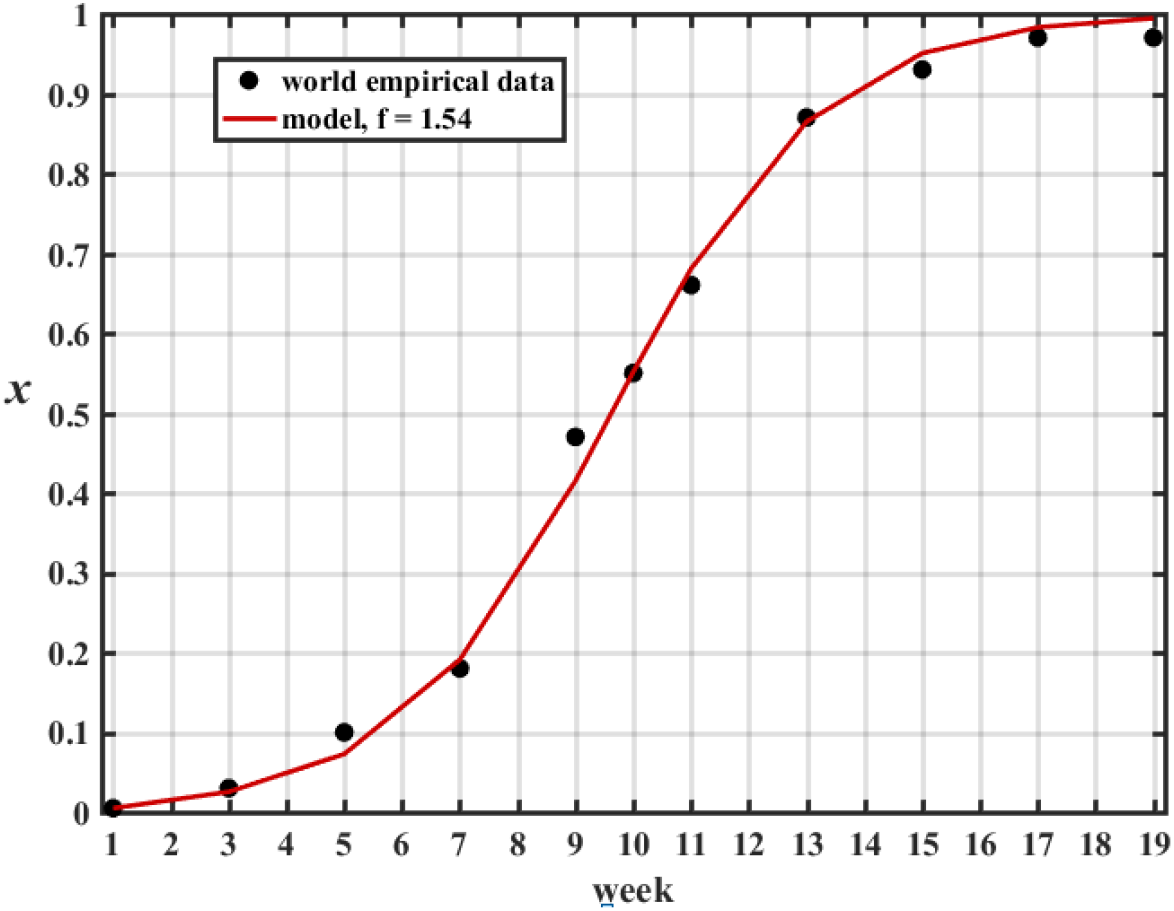
Change in the frequency of B.1.1.7. for the whole world: empirical data (black circles) and formula (4) (solid line).

## In summary

1. The change in frequency of SARS-CoV-2 virus lineage B1.1.7 in time is well described by formula (4) resulting from an elementary evolutionary model with a single parameter, the relative fitness *f*. This parameter can be interpreted as the relative fitness to the local wildtype.
2. The value of this parameter *f* slightly varies around 1.50 for most of the time series considered in this study: three of the four countries (Denmark, Switzerland and UK), the whole world and all of the five regions of Denmark except one (Nordjylland). This would imply that the variant is 50% more transmissible than the local wild type.
3. In the case of Netherlands, *f* is smaller (*f* = 1.33). One possible explanation is that, before the frequency reaches a value close to its asymptote *x* = 1, the fit strongly depends on the last data point. And, this point often has incomplete data and may change as more sequences come (Hodcroft 2021). Thus one thing to check is whether the fitness parameter converges to values closer to 1.5 when data for subsequent weeks in Netherlands is available. The data for Nordjylland, in turn, exhibits a very ‘jagged’ frequency - this often indicates low sequencing numbers (Hodcroft 2021) and seems to be the case since this Danish region is the one with lowest statistics of viral sequences.
4. As Hodcroft (2021) stressed, since in many countries sampling may not be equal across the country: samples may only cover one area or certain areas, it’s important not to assume frequencies shown are necessarily representative of the country. However, in the case of Denmark it seems there are no important variations across the different regions.

## Data Availability

All data available

https://outbreak.info/situation-reports?pango=B.1.1.7&loc=GBR&loc=USA&loc=USA_US-CA&selected=GBR

https://www.cdc.gov/coronavirus/2019-ncov/science/science-briefs/scientific-brief-emerging-variants.html?CDC_AA_refVal=https%3A%2F%2Fwww.cdc.gov%2Fcoronavirus%2F2019-ncov%2Fmore%2Fscience-and-research%2Fscientific-brief-emerging-variants.html

https://www.covid19genomics.dk/statistics

https://www.medrxiv.org/content/10.1101/2020.12.24.20248822v3

https://covariants.org

https://ispmbern.github.io/covid-19/variants/

## Acknowledgements

I Thank Raúl Donangelo and Edgardo García-Álvarez for their comments on this analysis. I am also grateful to Jorge Pullin, who drew my attention to the issue of fixation of the British variant of concern.

## Footnotes

1 Danish Covid-19 2021

2 Hodcroft 2021

